# Transcutaneous vagus nerve stimulation enhances reward-effort efficiency in major depressive disorder

**DOI:** 10.64898/2026.04.16.26351003

**Authors:** Paul A. G. Forbes, Emily Brandt, Mareike Aichholzer, Carmen Uckermark, Aicha Bouzouina, Leona Jacobsen, Jonathan Repple, Jonathan Kingslake, Christine Reif-Leonhard, Andreas Reif, Carmen Schiweck, Sharmili Edwin Thanarajah

**Affiliations:** Department of Psychiatry, Psychosomatic Medicine and Psychotherapy, University Hospital, Goethe University Frankfurt, Frankfurt am Main, Germany; Cooperative Brain Imaging Center (CoBIC), Goethe University Frankfurt am Main, Frankfurt, Germany; Institute for Translational Psychiatry, University of Münster, Münster, Germany; P1vital Products Limited, Howbery Park, Wallingford, Oxfordshire, UK; Fraunhofer Institute for Translational Medicine and Pharmacology, Frankfurt am Main, Germany

**Keywords:** major depressive disorder, vagus nerve stimulation, effort, reward

## Abstract

Major depressive disorder (MDD) is a highly prevalent psychiatric disorder with changes in motivation to work for rewards being a core symptom. Transcutaneous vagus nerve stimulation (tVNS) has emerged as a promising therapy but its effects on the core features of MDD, such as changes in motivation, remained relatively unexplored. In this randomised, single-blind, cross-over, controlled trial, we used a grip strength effort task to investigate how tVNS impacted choices to exert different levels of physical effort for varying monetary rewards in MDD patients (n=53) and a non-depressed control group (n=45). Compared to sham stimulation, tVNS enhanced the efficiency with which participants with severe depressive symptoms allocated physical effort for rewards (reward-effort efficiency). These effects were not seen in participants with less severe symptoms. Specifically, we found that the effect of tVNS on reward-effort efficiency was driven by reduced unnecessary effort, i.e., a reduction in choices to exert additional effort when this was not required to gain a larger reward. These findings suggest a potential motivational mechanism by which tVNS exerts its therapeutic effects in MDD. Determining whether the effects of tVNS are linked to broader changes in executive functioning, such as improvements in cognitive flexibility in MDD, should be a key aim for future work.

## Introduction

Major depressive disorder (MDD) is associated with anhedonia, including changes in motivation to gain rewards (Marx et al., 2023). Global cases of MDD have risen rapidly in the past decades (Yan et al., 2024) yet around one third of MDD patients do not respond to first-line treatment (Al-harbi, 2012; De Carlo et al., 2016). Therefore, new therapies, and an understanding of their potential mechanisms, are urgently needed to improve outcomes.

The vagus nerve originates in the brainstem and innervates major organs including the heart, lungs and gastrointestinal tract. It drives parasympathetic activity - slowing heart rate and breathing whilst promoting digestion - and has connections to brain circuits implicated in MDD (Howland, 2014), including noradrenergic, dopaminergic and serotonergic pathways (Brougher et al., 2021; Dorr & Debonnel, 2006; Han et al., 2018; Onimus et al., 2026; Ruffoli et al., 2011). Surgically implanted vagus nerve stimulators act on the body-brain axis and are an approved therapy for treatment-resistant MDD. Despite having received increased attention in the past two decades (Edwin Thanarajah & Reif, 2022), the underlying mechanisms of vague nerve stimulation are yet to be fully elucidated and have been difficult to explore due to the invasive nature of the procedure. However, the advent of non-invasive transcutaneous vagus nerve stimulators has enabled the investigation of the underlying mechanisms.

Transcutaneous vagus nerve stimulation (tVNS) has shown promise as a non-invasive treatment for MDD (for reviews, see Edwin Thanarajah & Reif, 2022; Guerriero et al., 2021; Cimpianu et al., 2017; Parente et al. 2024; Müller et al., 2018). Several randomised controlled trials (Hein et al., 2013; Li et al., 2022) and cohort studies (Rong et al., 2016) have shown improvements in MDD symptoms following multiple weeks of tVNS. Results from these non-invasive tVNS studies support findings showing the long-term beneficial effects of invasive implanted vagus nerve stimulators in treatment-resistant MDD (Bajbouj et al., 2010). In the brain, tVNS in MDD could increase functional connectivity between amygdala and prefrontal cortex (Liu et al., 2016) and alter default mode network connectivity (Fang et al., 2016). In terms of psychological effects (for a review, see Colzato & Beste, 2020), MDD patients showed enhanced positive emotion recognition following short-term tVNS (Zhao et al., 2025) and a decrease in the recognition of sad emotions (Koenig et al., 2021).

As mentioned above, whilst MDD is associated with changes in emotional processing, it is also defined by anhedonia (American Psychiatric Association et al., 2013), yet the effects of tVNS on motivation and reward processing in MDD remain relatively unexplored. Impaired motivation in MDD is an important treatment target (Serretti, 2025; Valton et al., 2025) as anhedonia is a key negative predictor of recovery in MDD (McMakin et al., 2012). Rodent studies indicate that both electrical and optogenetic activation of the vagus nerve modulate activity in dopaminergic pathways critical for motivation (Brougher et al., 2021; Choi et al., 2024; Han et al., 2018; Surowka et al., 2015). Dopaminergic activity is strongly implicated in both reward and effort processing (Castrellon et al., 2021; Mkrtchian et al., 2025; Soutschek et al., 2023): methylphenidate enhances willingness to exert cognitive effort by modulating striatal dopaminergic signalling (Westbrook et al., 2020), and dopaminergic neurons encoded both expected rewards and anticipated effort costs (Varazzani et al., 2015). Thus, tVNS could alter choices to exert effort for rewards in MDD via its impact on cost-benefit dopaminergic signalling.

Moreover, VNS could influence noradrenaline activity in the locus coeruleus (Dorr & Debonnel, 2006; Ruffoli et al., 2011; Schneider et al., 2025; Su et al., 2025) and activity in this region plays a critical role in dynamically tracking the amount of effort produced to tackle cognitive and physical challenges (Borderies et al., 2020; Bornert & Bouret, 2021).

To date, tVNS studies investigating effort-related behaviour in MDD have focused on the process of effort exertion rather than on *decisions* to exert effort or not. Decisions to exert effort for a reward involve a cost-benefit calculation (“*Is this reward worth the effort?”*) between the potential rewards on offer and the required effort to obtain them - as effort costs increase the value of rewards typically decreases (Chong et al., 2017; Robbins & Everitt, 1996). Conversely, studies investigating the process of effort exertion track ongoing motivation once the decision to exert effort has already been made. For example, Ferstl et al. (2024) investigated whether short-term tVNS increased motivation during effort exertion (button presses) in MDD patients and control participants who completed a tVNS session and sham session in a counterbalanced order. The key measures were effort maintenance (frequency of button presses) and effort invigoration (the initial increase in the frequency of button presses). When analysing the MDD and control participants together, tVNS led to greater effort invigoration in the first session (there was no effect when looking at both sessions), thereby partially replicating their previous work in healthy participants (Neuser et al., 2020). When Lucchi et al. (2024) conducted a direct replication of Neuser et al.’s study, they did not find evidence that tVNS increased effort invigoration (or effort maintenance). Thus, whilst one study hints that tVNS could alter the process of effort-related behaviour in MDD, the evidence remains inconclusive.

In the present study, rather than focus on the process of effort exertion (i.e., invigoration and maintenance), we investigated how tVNS impacted effort-related *decision-making* in MDD. Several studies have shown improvements in decision making following vagus nerve stimulation in rats (Cao et al., 2016) and healthy humans (Keute et al., 2020; Su et al., 2025). Here, our specific focus centred on *reward-effort efficiency* which captured how optimally individuals decided to allocate effort to gain rewards. An efficient agent exerts greater effort only when this is necessary to obtain a larger reward, but refrains from engaging in unnecessary effort, thereby maximising the cost-benefit ratio. Thus, an *inefficient* agent (1) chooses to exert greater effort even when this does not lead to a larger reward (*enhanced unnecessary effort*) and/or (2) chooses not to exert greater effort when this could lead to a larger reward (*reduced necessary effort*).

Most MDD studies have focused on *reduced necessary effort*: when offered the opportunity to gain larger rewards, MDD patients are less willing to engage in more complex working memory tasks (Ang et al., 2023; Westbrook et al., 2023), make more numeric decisions (Hershenberg et al., 2016), perform more button presses (Berwian et al., 2020; Treadway et al., 2012; Yang et al., 2014; Zou et al., 2020) and exert more force when squeezing a hand dynamometer (Cléry-Melin et al., 2011; Horne et al., 2021; Malik et al., 2025; Valton et al., 2025). Although not all studies show a reduced willingness to exert effort for rewards in MDD (Barch et al., 2023; Bustamante et al., 2024; Cathomas et al., 2021; Culbreth et al., 2024; Yang et al., 2021), a recent meta-analysis of 23 effects showed a small negative effect of MDD (g = -.20) versus controls (Pillny et al., 2024).

Yet, *enhanced unnecessary effort* remains relatively unexplored in MDD. This is surprising given that an imbalance between the effort we exert and the rewards we receive has been linked to life satisfaction (Braunheim et al., 2024). Additionally, effort-related choices in everyday life do not simply involve exerting more effort indiscriminately. Instead, we must decide how to allocate our limited physical and cognitive resources to maximise potential outcomes given our current motivational state (Hewitt et al., 2025). We evaluate which options are *worth the effort* and which are not. Furthermore, engaging in unnecessary effort could exacerbate fatigue symptoms in MDD and Steward et al. (2025) showed that fatigue predicted increased perceived effort costs in MDD. In their meta-analysis, Pillny et al. (2024) reported a ‘maladaptive effort/reward computation’ in MDD patients - decisions to exert effort were less influenced by the magnitude and probability of a potential reward. In a recent study of 16-25 year olds, Sahni et al. (2025) showed that as anticipatory anhedonia increased the difference in the effort exerted for low versus high rewards decreased. In other words, those with more anhedonia symptoms did not modulate the effort they exerted as a function of reward value. This hints at less efficient decision-making in MDD in terms of maximising potential rewards relative to exerted effort.

Given initial reports of improvements in effort-related behaviour in MDD following tVNS (Ferstl et al., 2024) and enhanced decision-making following VNS (Cao et al., 2016; Keute et al., 2020; Su et al., 2025), we hypothesised that tVNS would improve reward-effort efficiency by reducing *unnecessary effort* and/or increasing *necessary effort*. Crucially, we aimed to determine whether a diagnosis of MDD and depressive symptom severity modulated potential tVNS effects.

## Methods

### Study design and procedure

The study was conducted as part of the clinical trial “MODULATE Depression” (German Clinical Trials Register ID: DRKS00024823), a randomized, single-blind design approved by the Ethics Committee of the Goethe University Frankfurt (Approval ID: 2021-48). The study followed a within-subject crossover design, consisting of one screening visit and two test days. On the two test days, participants received transcutaneous auricular vagus nerve stimulation (tVNS) and sham stimulation in counterbalanced order, with stimulation order assigned according to a predefined randomisation list. Both test days followed the same protocol, with the stimulation condition (tVNS or sham) being the only difference. Test days one and two were scheduled within a maximum of ten days to reduce relevant variability in symptom severity.

Eligibility was verified during a screening visit. Current and lifetime psychiatric disorders were assessed using the Mini-International Neuropsychiatric Interview (MINI) 7.0.2 to identify potential comorbid psychiatric diagnoses meeting exclusion criteria (Sheehan et al., 1998). The Montgomery-Asberg Depression Rating Scale (MADRS) was used to assess depressive symptom severity via a clinical interview (Montgomery & Asberg, 1979) and self-reported depressive symptom severity was assessed with the Beck Depression Inventory–II (BDI-II) (Beck et al., 1996). Additionally, the Perceived Stress Scale (PSS-10) was used to assess perceived stress in the previous four weeks (Cohen, 1988; Cohen et al., 1983) and the Childhood Trauma Questionnaire was also collected (Bernstein et al., 1994). Questionnaire scores as well as anthropometric and demographic information are presented in Table 1.

**Table 1.**
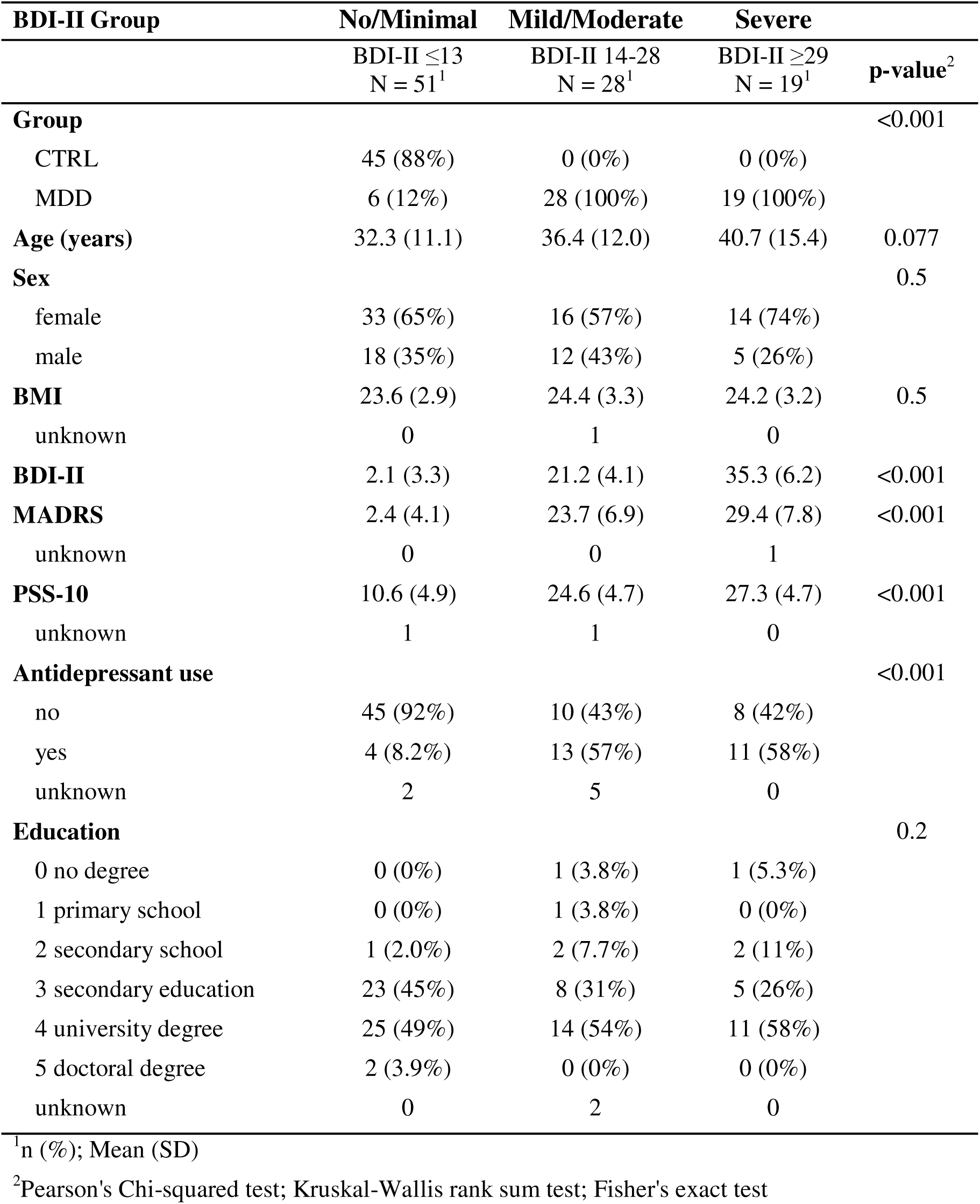
A comparison of the participants in the three groups based on BDI-II scores. The BDI-II score was collected on three occasions (screening, T1, T2). The scores in this table are the scores from the screening date for 96 participants. Two participants did not provide a BDI-II score at screening so we included their BDI-II score from T1. This table was created using the *gtsummary* package in R (Sjoberg et al., 2021).

| BDI-II Group | No/Minimal | Mild/Moderate | Severe |  |
| --- | --- | --- | --- | --- |
| | BDI-II $\leq 13$<br>N = 51<sup>1</sup> | BDI-II 14-28<br>N = 28<sup>1</sup> | BDI-II $\geq 29$<br>N = 19<sup>1</sup> | p-value<sup>2</sup> |
| **Group** |  |  |  | <0.001 |
| CTRL | 45 (88%) | 0 (0%) | 0 (0%) |  |
| MDD | 6 (12%) | 28 (100%) | 19 (100%) |  |
| **Age (years)** | 32.3 (11.1) | 36.4 (12.0) | 40.7 (15.4) | 0.077 |
| **Sex** |  |  |  | 0.5 |
| female | 33 (65%) | 16 (57%) | 14 (74%) |  |
| male | 18 (35%) | 12 (43%) | 5 (26%) |  |
| **BMI** | 23.6 (2.9) | 24.4 (3.3) | 24.2 (3.2) | 0.5 |
| unknown | 0 | 1 | 0 |  |
| **BDI-II** | 2.1 (3.3) | 21.2 (4.1) | 35.3 (6.2) | <0.001 |
| **MADRS** | 2.4 (4.1) | 23.7 (6.9) | 29.4 (7.8) | <0.001 |
| unknown | 0 | 0 | 1 |  |
| **PSS-10** | 10.6 (4.9) | 24.6 (4.7) | 27.3 (4.7) | <0.001 |
| unknown | 1 | 1 | 0 |  |
| **Antidepressant use** |  |  |  | <0.001 |
| no | 45 (92%) | 10 (43%) | 8 (42%) |  |
| yes | 4 (8.2%) | 13 (57%) | 11 (58%) |  |
| unknown | 2 | 5 | 0 |  |
| **Education** |  |  |  | 0.2 |
| 0 no degree | 0 (0%) | 1 (3.8%) | 1 (5.3%) |  |
| 1 primary school | 0 (0%) | 1 (3.8%) | 0 (0%) |  |
| 2 secondary school | 1 (2.0%) | 2 (7.7%) | 2 (11%) |  |
| 3 secondary education | 23 (45%) | 8 (31%) | 5 (26%) |  |
| 4 university degree | 25 (49%) | 14 (54%) | 11 (58%) |  |
| 5 doctoral degree | 2 (3.9%) | 0 (0%) | 0 (0%) |  |
| unknown | 0 | 2 | 0 |  |

<sup>1</sup>n (%); Mean (SD)
<sup>2</sup>Pearson's Chi-squared test; Kruskal-Wallis rank sum test; Fisher's exact test

After the screening visit, eligible participants were assigned to tVNS and sham stimulation for the following test days (Figure 1). The study was single-blind, with participants blinded to stimulation conditions.

**Figure 1.**
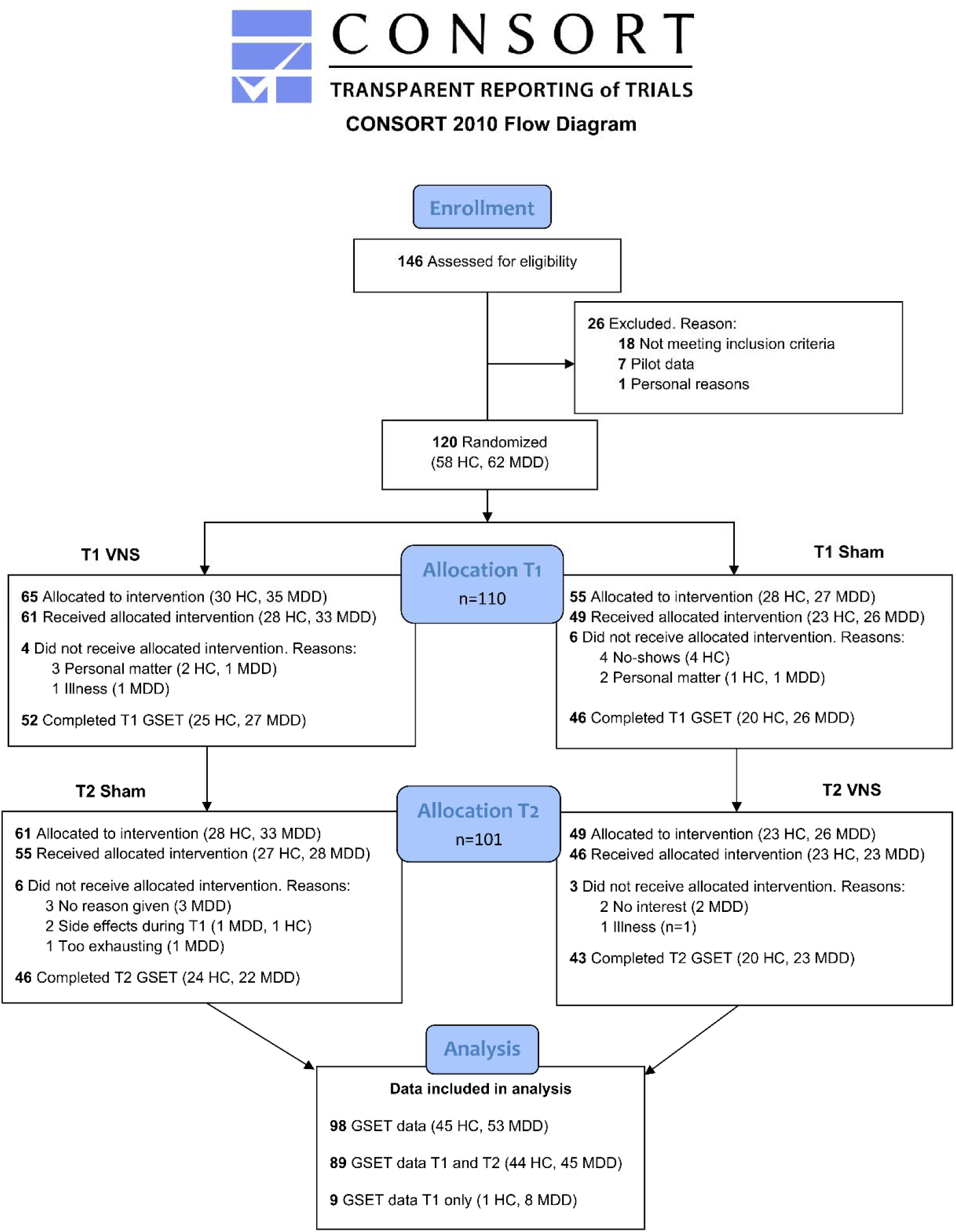
Consort Flow Diagram of the randomised controlled trial (MODULATE study). The analysis comprised participants who received the allocated intervention and completed the GSET assessment at least once (n = 98), with reasons for exclusions and drop-outs detailed at each stage. The GSET assessment was implemented following a protocol amendment. As a result, participants randomised prior to this amendment were not eligible for inclusion in the GSET analysis.

Test days were scheduled within 10 days to reduce variability in symptom severity. Participants were invited in the morning after an overnight fasting period of at least 8 hours. At the beginning of each test day, blood samples were collected. Participants then completed the BDI-II to assess the severity of current depressive symptoms (Beck et al., 1996) and completed visual analogue scales to assess their current positive and negative emotions. Stimulation (tVNS or sham stimulation) was initiated and commenced for approximately 10 minutes prior to the first behavioural task.

Participants completed three behavioural computer tasks in a fixed sequence: the Grip Strength Effort Task (GSET, the task analysed in this paper), a modified version of the Montreal Stress Imaging Task (MIST) integrating elements of Dedovic et al. (2005), followed by a reinforcement learning task. Only the results from the GSET are reported in the current manuscript (results of the other tasks can be found in Schiweck et al., 2025 and Zhao et al., 2025). Continuous electrocardiography was recorded during the modified MIST and saliva samples were collected at several time points during the sessions (both not analysed here)

### Participants

Participants were recruited between May 2021 and July 2023 from the Department of Psychiatry, Psychosomatics and Psychotherapy of the University Hospital Frankfurt. All participants were aged 18-65 years and provided written informed consent. Participants in the MDD group met DSM-5 criteria for a current unipolar major depressive episode. Exclusion criteria for the MDD group included bipolar disorder, dementia, psychotic disorders and substance use disorder, as well as substance- or medication-induced depressive disorder. Healthy controls were required to have no current or past depressive episode. For both groups, exclusion criteria included body mass index (BMI) > 35kg/m2, autoimmune, rheumatological, neurological or metabolic diseases. Pregnancy, lactation or an acute infection at the time of assessment were also defined as exclusion criteria. An overview of the participant allocation and exclusions are provided in Figure 1.

### Transcutaneous auricular vagus nerve stimulation

Transcutaneous auricular vagus nerve stimulation (tVNS) was applied using the NEMOS® device (tVNS Technologies, Erlangen, Germany). For verum stimulation, electrodes were placed at the cymba conchae of the right ear, targeting the auricular branch of the vagus nerve. Sham stimulation was applied at the right earlobe (Figure 2A).

**Figure 2.**
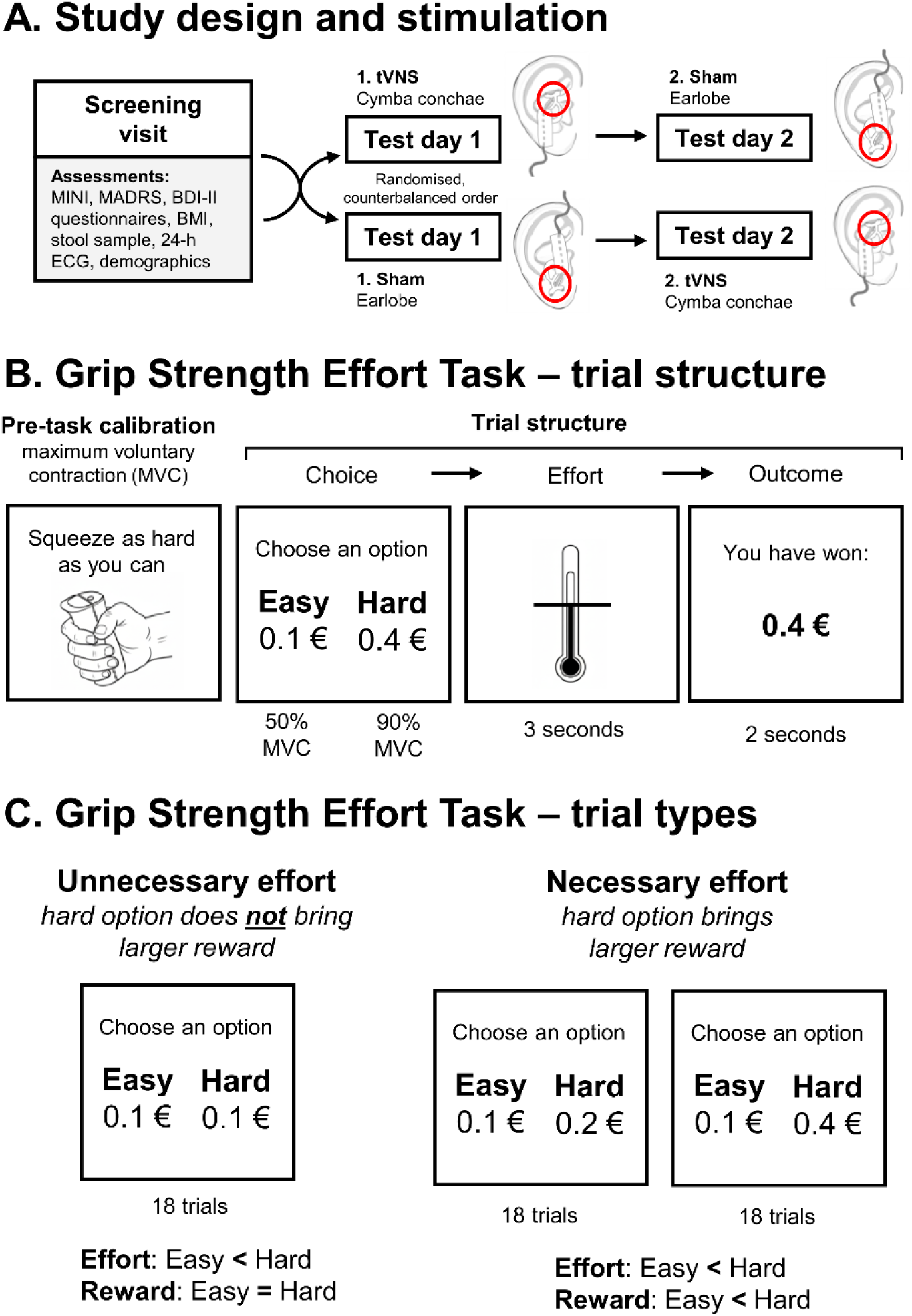
**A: Study design and stimulation.** Following a screening visit (2 h), participants attended two test days (2.5 h each). In a randomised, single-blind, crossover design, they received tVNS at the cymba conchae or sham stimulation at the earlobe of the right ear on separate test days. **B: Trial structure of the Grip Strength Effort Task (GSET).** After calibration of maximum voluntary contraction (MVC) with the dominant hand, participants chose between an Easy (50% of the MVC) and a Hard option (90% of the MVC) on each trial. Following a 2 s preparation phase (“Ready?”; not shown), participants were able to exert force for 3 s while receiving real-time visual feedback. The reward outcome was presented afterwards for 2 s. **C: Trial types in the GSET.** The task included three effort-reward combinations each presented 18 times (54 trials in total): an unnecessary effort condition (0.1€-Easy vs. 0.1€-Hard) and two necessary effort conditions (0.1€-Easy vs. 0.2€-Hard or 0.4€-Hard).

On each test day, the stimulation was started after blood sampling and questionnaire completion and was started approximately ten minutes prior to the first task. The stimulation was applied continuously throughout the test days and lasted for approximately 90 minutes in each session. Stimulation was applied in biphasic electrical pulses with a pulse width of 250 µs and a frequency of 25 Hz, with alternating periods of stimulation and pause (32 s on / 28-30 s off). To ensure stable contact, electrode pads and conductive gel were used. Stimulation intensity was individually determined at the start of each test day. The current started at 0.1 mA and was gradually increased in steps of 0.1 mA until participants reported an uncomfortable tingling sensation at the stimulation site, then decreased until the sensation was clearly perceptible but did not feel painful. The individual threshold and chosen stimulation intensity were documented for each participant and session. The maximum stimulation intensity was 5 mA, median stimulation intensity was approximately 0.80-0.85 mA.

Right-sided stimulation was chosen due to evidence suggesting stronger functional connectivity of the right vagus nerve to limbic and dopaminergic brain circuits involved in reward and emotional processing (Brougher et al., 2021; Han et al., 2018; Müller et al., 2022).

The blinding was successful as participants could not reliably distinguish between the actual and sham stimulation. Of the 85 participants who completed both sessions, 71 participants indicated their belief in the treatment condition for both sessions, i.e. whether they thought they had received actual tVNS or sham stimulation. Only 11 of these participants (15.5%) correctly identified when they received sham stimulation and when they received actual tVNS, which is below the 25% expected by chance.

### Grip strength effort task (GSET) - measuring reward-effort efficiency

The Grip Strength Effort Task (GSET) assessed the willingness to exert physical effort in exchange for monetary rewards using a hand grip dynamometer (Kinvent Biomécanique, Montpellier, France) which recorded grip strength in real time (Figure 2B; Figure 2C). The task was delivered via the P1vital ePRO Clinical System (P1vital Products Limited, Wallingford, UK)

Before the main task, participants completed four calibration trials (two per hand) to determine their maximum voluntary contraction (MVC; each participant’s individual maximum grip strength). The task was then performed with the dominant hand.

The main task compromised 54 trials. On each trial, participants chose between an ‘Easy’ and a ‘Hard’ option. The Easy option was the same on each trial - participants received 0.1 € if they successfully squeezed the hand dynamometer to 50% of their MVC. There were three different reward levels when choosing the Hard option: participants could receive either 0.1, 0.2 or 0.4 € and had to squeeze the hand dynamometer at 90% of their MVC. Thus, participants faced three different combinations of Easy and Hard options which they were presented 18 times each, resulting in 54 trials overall (Figure 2).

At the beginning of each trial the potential monetary reward was displayed and participants then had to select either the Easy or Hard option. A 3-second period followed during which they had to exert the required force. A bar was displayed on screen to provide real-time feedback concerning the applied force. If the applied force exceeded the threshold (50% for the Easy option and 90% for the Hard option), the trial was successful and they received the reward. Participants received immediate feedback as to whether they exerted sufficient effort to receive the reward.

To quantify whether participants maximised reward relative to exerted effort, the primary outcome was *reward-effort efficiency*: this was coded as a 1 when participants chose the Easy option when both rewards were equal (0.1€-Easy [50% MVC] vs. 0.1€-Hard [90% MVC]) or when they chose the Hard option when this choice offered a larger reward (0.1€-Easy vs. 0.2€-Hard; 0.1€-Easy vs. 0.4€-Hard; Figure 2C). Conversely, reward-effort efficiency was coded as a 0 when participants exerted unnecessary effort (choosing the Hard option when both rewards were equal) or when they chose not to exert additional effort to receive a larger reward (choosing the Easy option despite this leading to a smaller reward).

## Statistical analysis

We conducted a mixed effects logistic regression using the R package *glmmTMB* (McGillycuddy et al., 2025). To determine the statistical significance of fixed effects, we performed omnibus Type-III Wald χ2 tests using the R package *car* (Fox et al., 2026). The key dependent variable was *reward-effort efficiency* which was coded as either a 0 or 1 depending on whether a participant’s choice aimed to maximise their rewards relative to effort required to obtain the reward.

To probe the effect of tVNS on reward-effort efficiency, we split the data according to unnecessary effort (i.e., choosing the Hard option when the Easy option was available despite equal rewards) and necessary effort (i.e., choosing the Hard option when it resulted in a larger reward) (see Figure 2C).

For unnecessary effort, we analysed trials where the lowest reward (0.1€) was on offer either for 50% or 90% effort (Easy vs. Hard). We coded a variable, *unnecessary effort*, which equalled a 0 when participants chose the 0.1€-Easy (50% MVC) option and a 1 when participants chose the 0.1€-Hard (90% MVC) option (Figure 2C).

For necessary effort, we analysed trials where a larger reward was on offer for greater effort (0.1€-Easy vs. 0.2€-Hard; 0.1€-Easy vs. 0.4€-Hard). We coded a variable, *necessary effort*, which equalled a 0 when participants chose the 0.1-Easy option and a 1 when participants chose the higher effort, higher reward option (either 0.2-Hard or 0.4-Hard).

To determine whether the effects of tVNS were driven by depressive symptom severity, we coded a factor BDI-II group by splitting participants into three groups (see Table 1) based on established BDI-II cut offs: no-to-minimal symptoms (≤13; n=51; reference group; mainly healthy controls), mild-to-moderate symptoms (14-28; n=28), or severe symptoms (≥29; n=19). All participants in the mild to moderate and severe BDI-II groups had a MDD diagnosis, whereas the no-to-minimal group was largely made up of healthy control participants (88%) with just six MDD patients in this group (Table 1). We repeated all analyses using BDI-II scores as a continuous variable.

In the models, we included the key interaction between condition (sham, tVNS) and group (Control, MDD) or condition (sham, tVNS) and BDI-II group (no-to-minimal, mild-to-moderate, severe) and, as control variables, test day (1, 2), trial number (centred), age (centred) and sex (female, male). We included random intercepts for participants and random slopes for the effect of condition and trial number (centred). Trials in which participants did not squeeze to the required effort level (0.73% of all trials) were excluded from the analysis.

## Results

### tVNS enhances reward-effort efficiency in severe MDD

There was no interaction between group (Control vs. MDD group) and condition (type III Wald test, χ2[1] = 2.32, p=0.128) for reward effort-efficiency. When we repeated the analysis replacing group (Control, MDD) with BDI-II group (no-to-minimal, mild-to-moderate, severe), there was a significant interaction between BDI-II group and condition (χ2[2] = 6.34, p=0.042) driven by the severe BDI-II group (β=0.931, SE=0.370, p=0.012) (Figure 3A). Whilst tVNS resulted in a significant increase in reward-effort efficiency in the severe BDI-II group (mean [SE] estimated probabilities: sham=0.894 [0.034], tVNS=0.946 [0.021]; p=0.022), there were no effects of tVNS on reward-effort efficiency in the no-to-minimal BDI-II group (sham=0.950 [0.012], tVNS=0.940 [0.015]; p=0.360) or mild-to-moderate BDI-II group (sham=0.940 [0.017], tVNS=0.941 [0.019] ; p=0.939). Note, when we treated BDI-II score as a continuous variable and repeated the above analysis, there was also a significant interaction between BDI scores and condition (χ2[1] = 5.42, p=0.020, β=0.024, SE=0.010) thereby replicating the effect of the categorical BDI group analysis.

**Figure 3.**
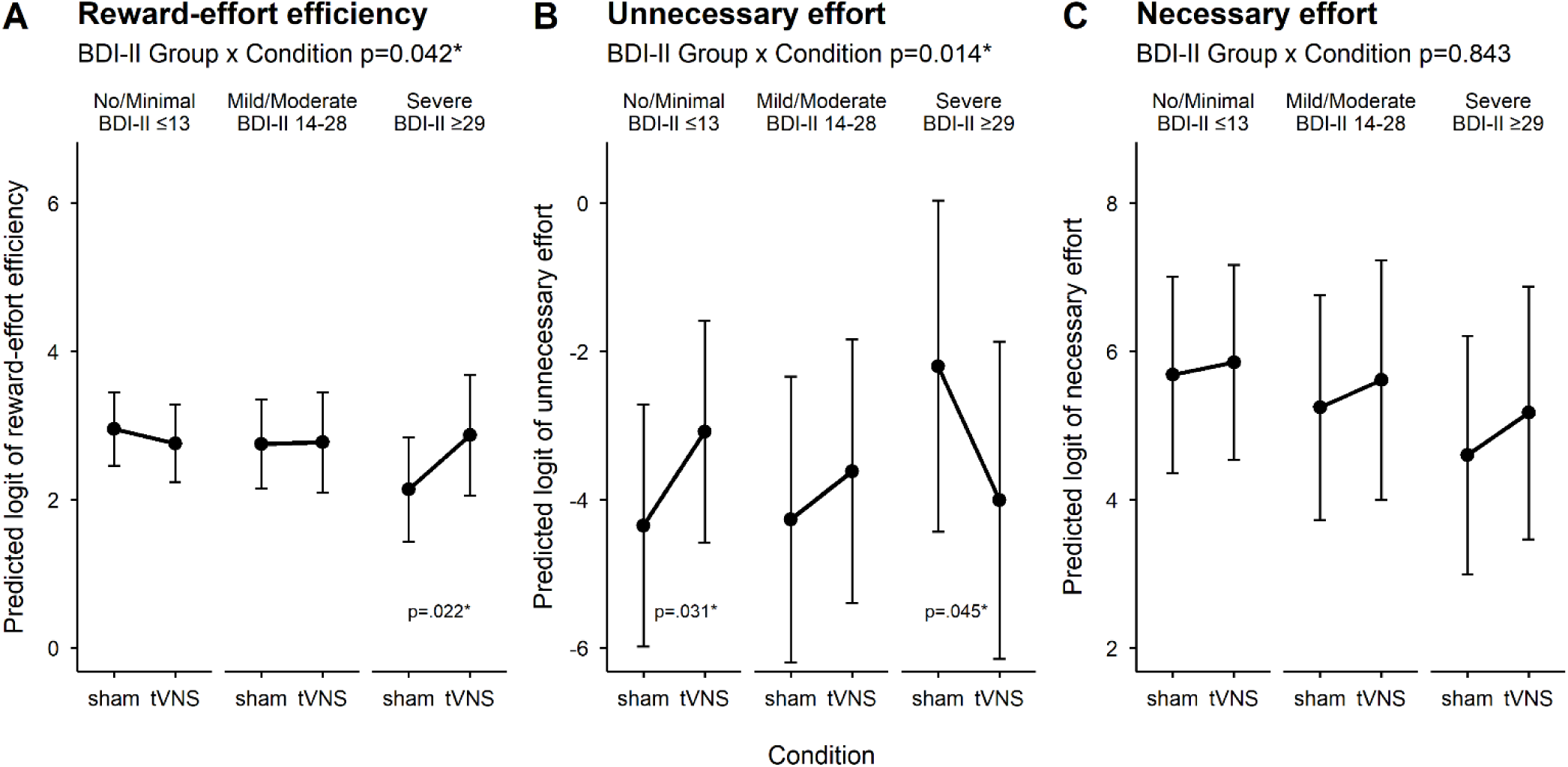
tVNS effects on reward-effort choices across depressive symptom severity. **A**: tVNS enhanced reward-effort efficiency (log odds of maximising reward relative to effort) in participants with more severe depressive symptoms (BDI-II score ≥ 29). **B**: Follow up analysis showed that increases in reward-effort efficiency under tVNS were primarily driven by reductions in unnecessary effort in those with severe depressive symptoms. Conversely, those in the no-to-minimal BDI-II group showed an increase in unnecessary effort under tVNS compared to sham. **C:** There was no significant interaction between BDI-II and condition for necessary effort. Estimated marginal means are shown for each condition and BDI-II group extracted from the mixed effects models using the R package *emmeans* (Lenth, 2022); error bars represent the asymptotic 95% CI.

### tVNS effects on reward-effort efficiency driven by reductions in unnecessary effort in severe MDD

The key interaction between BDI-II group and condition was significant (χ2[2] = 8.56, p=0.014) (Figure 3B). Specifically, the interaction term for the severe BDI-II group, compared to the no-to-minimal group, was significant (β=-3.08, SE=1.05, p=0.004). Whilst tVNS resulted in an *increase* in unnecessary effort in the no-to-minimal BDI-II group (mean [SE] estimated probabilities: sham=0.013 [0.011], tVNS=0.044 [0.032]; p=0.031), there was a *decrease* in unnecessary effort in the tVNS condition compared to the sham condition in the severe BDI-II group (sham=0.100 [0.102], tVNS=0.018 [0.020]; p=0.045). There were no effects of tVNS on unnecessary effort in the mild-to-moderate BDI-II group (sham= 0.014 [0.013], tVNS=0.026 [0.023] ; p=0.418). As above, when we treated BDI-II scores as a continuous variable and repeated the analysis, there was a significant interaction between BDI-II scores and condition for unnecessary effort (χ2[1] = 4.03, p=0.045, β=-0.057, SE=0.029)

For necessary effort, the key interaction between BDI-II group and condition was not significant (χ2[2] = 0.342, p=0.843) with no differences in necessary effort across the sham and tVNS conditions in any of the BDI-II groups (Figure 3C). Similarly, when we treated BDI-II scores as a continuous variable, there was no significant interaction between BDI-II scores and condition for necessary effort (χ2[1] = 1.49, p=0.222).

Taken together the results suggested that the increases in reward-effort efficiency seen in participants with more severe depressive symptoms (severe BDI-II group) was driven more by decreases in unnecessary effort, rather than increases in necessary effort.

## Discussion

We investigated how short-term tVNS impacted reward-effort efficiency in 98 participants (45 control; 53 MDD). The effect of tVNS on reward-effort efficiency was dependent on MDD symptom severity. Specifically, compared to sham, tVNS enhanced reward-effort efficiency in those participants with severe depressive symptoms (BDI-II scores ≥29). These effects were not present in participants with no or more moderate depressive symptoms where tVNS did not alter reward-effort efficiency. When we compared the effect of tVNS on *necessary* effort (choosing to exert more effort for larger rewards) with *unnecessary* effort (choosing to exert more effort even though this does not lead to a larger reward), we found that the effect of tVNS on reward-effort efficiency was driven by reductions in unnecessary effort in participants with severe depressive symptoms. These findings suggest that tVNS leads to a more efficient allocation of effort when gaining rewards in MDD and suggest a potential motivational mechanism by which tVNS exerts its therapeutic effects.

Our findings support previous work showing changes in effort-based decision making in MDD (for a recent meta-analysis, see Pillny et al., 2024). Whilst previous work has focused on MDD patients’ reduced willingness to gain larger rewards for more effort (reduced necessary effort), we found differences in reward-effort efficiency - MDD patients with more severe symptoms chose to exert effort even though this was not necessary to gain a larger reward (enhanced unnecessary effort). Notably, a single-session of tVNS improved reward-effort efficiency and led to decision making comparable to non-depressed control participants and patients with less severe depressive symptoms (Figure 3).

Previous work investigating the modulation of effort-related behaviour using tVNS, has focused on the process of effort exertion, specifically, the frequency of button presses, and found inconsistent effects (Ferstl et al., 2024; Lucchi et al., 2024; Neuser et al., 2020). Here, we show for the first time that tVNS can lead to improvements in effort-related decision-making in MDD. Our findings do not support previous studies showing general improvements in decision making following VNS (Cao et al., 2016; Keute et al., 2020), instead the beneficial effects of tVNS were dependent on the severity of depressive symptoms. This suggests that tVNS may be most effective in instances where there is greater potential for improved performance. Reward-effort efficiency was approximately 95% in the control group under sham stimulation, thus there was limited potential for tVNS to improve performance. In contrast to participants with severe MDD symptoms, those with no or minimal MDD symptoms showed an *increase* in unnecessary effort under tVNS, i.e., a detrimental effect of stimulation on performance (Figure 3B). Su et al. (2025) found the greatest improvements in a perceptual decision-making task following tVNS in less proficient participants and “only when additional cognitive resources were required” (p. 984). Similarly, An et al. (2025) found that only participants with low baseline performance in working memory tasks improved following tVNS. We have found analogous findings in terms of the physiological effects of tVNS - stimulation improved cardiac vagal reactivity to stress but only in those participants with baseline low heart rate variability (Schiweck et al., 2025). Thus, tVNS could help to normalise suboptimal cognitive and physiological responses but provide limited benefit (or even detrimental effects) when responses are already at or near optimal.

Efficient performance in the grip strength effort task involved adaptively switching between Easy and Hard levels of effort depending on the reward level. Previous work investigating executive functioning in MDD has reported consistent deficits in switching, for example, between task sets or response rules (for a meta-analysis, see Snyder, 2013). In healthy participants, tVNS has been shown to increase cognitive flexibility in a set-shifting paradigm (Borges et al., 2020) and a meta-analysis suggested that tVNS may have its strongest cognitive effects on executive functioning (Ridgewell et al., 2021). Several studies have reported improved executive functioning following tVNS in MDD (Desbeaumes Jodoin et al., 2018; Sackeim et al., 2001). Thus, whether tVNS has a general effect on cognitive flexibility in MDD, rather than a specific effect on effort-related decision making, is an exciting avenue for future work. Moreover, future work will need to test alternative explanations for our findings beyond cognitive flexibility, such as changes in attention and perception following tVNS (Aniwattanapong et al., 2022; Su et al., 2025).

Stimulation of the vagus nerve results in a plethora of physiological changes (Howland, 2014; Ruffoli et al., 2011), thus the neural mechanisms by which tVNS modulates effort-related decision making in MDD is unclear. Noradrenergic activity has been implicated in effort-related decision making (Borderies et al., 2020; Bornert & Bouret, 2021) and animal work suggests VNS could exert its effects by increasing locus coeruleus noradrenergic activity (Dorr & Debonnel, 2006; Grimonprez et al., 2015). Yet, whether tVNS has similar noradrenergic effects in humans remains unclear (Burger et al., 2020). In a well-powered study using optimised stimulation parameters, D’Agostini et al. (2022) found no effect of tVNS on a range of noradrenergic markers in healthy participants, including pupil dilation, salivary alpha-amylase, respiratory rate, cardiac vagal modulation, and salivary flow rate. In a recent meta-analysis, Pervaz et al., (2025) found that only pulsed stimulation protocols, not conventional stimulation protocols as used in the present study, increased pupil size. Alternatively, tVNS could exert its effects in MDD via dopaminergic mechanisms which have also been implicated in effort-related behaviour (Michely et al., 2020). Work in rodents shows that stimulation of the vagus nerve activates the midbrain dopamine system (Brougher et al., 2021; Choi et al., 2024) and neuroimaging studies in MDD patients suggest modulation of the dopaminergic system following tVNS (Wang et al., 2018; Zhang et al., 2022). We intentionally stimulated the right vagus nerve to optimise the impact on the dopaminergic system (Han et al., 2018). Future work using carefully designed cognitive tasks combined with neuroimaging and psychopharmacology will be useful in elucidating the exact neurocomputational mechanisms of tVNS in MDD.

We split participants into three groups based on well-established cut offs for BDI-II scores. However, this left just 19 participants in the severe BDI-II group. Future studies investigating tVNS effects should aim to replicate our findings with larger samples to ensure the robustness of our effects in those with more severe depressive symptoms. It is worth noting, however, that when we treated BDI scores as a continuous rather than categorical variable, we replicated our key findings showing that tVNS effects on reward-effort efficiency and unnecessary effort were dependent on depressive symptom severity.

Changes in motivation in MDD have been difficult to target with existing therapies (Serretti, 2025; Valton et al., 2025), thus our findings provide initial evidence that tVNS could lead to changes in how effort is allocated to gain rewards in MDD. In the present study, we observed acute effects of tVNS in MDD patients, however, studies using invasive VNS have involved treatment-resistant MDD patients and studied outcomes over longer timescales. Future studies will need to establish the robustness of tVNS effects in different populations of MDD patients over longer timescales, that is, days and weeks rather than hours, and with different stimulation protocols (Pervaz et al., 2025). This is especially pertinent given the inconclusive evidence concerning the effects of tVNS on effort-related behaviour in healthy participants (cf. Lucchi et al., 2024; Neuser et al., 2020) and those with an MDD diagnosis (Ferstl et al., 2024).

To conclude, short-term tVNS improved reward-effort efficiency in patients with severe depressive symptoms. These effects were driven by reductions in unnecessary effort. Existing therapies have struggled to target motivational deficits in MDD. Crucially, therefore, our findings provide initial evidence that tVNS could optimise the allocation of effort to obtain rewards in MDD and highlight exciting avenues for future work. Firstly, we must establish how specific these tVNS effects are to reward-effort efficiency - it is possible that tVNS effects in MDD are better accounted for by a general enhancement of executive functions, such as cognitive flexibility. Secondly, tVNS results in a diverse range of poorly understood physiological changes - understanding their exact mechanism in MDD could help to reveal new treatment targets. Here, combining tVNS with psychopharmacological interventions and neuroimaging could be particularly fruitful.

## Data Availability

Data produced in the present study are available upon reasonable request to the authors

